# SPONTANEOUS ECHO CONTRAST IN MITRAL STENOSIS: WHY AND HOW?

**DOI:** 10.1101/2020.03.31.20046318

**Authors:** Ziya Gokalp Bilgel, Ibrahim Hakan Gullu, Saif Hamad, Mutlu Kasar, Tansel Erol, Senol Demircan, Haldun MUderrisoglu

**Affiliations:** Department of Cardiology, Faculty of Medicine, Baskent University, Adana, Turkey; Diskapi Training and Research Hospital, Ankara, Turkey; Department of Hematology, Faculty of Medicine, Baskent University, Adana, Turkey; Department of Cardiology, Faculty of Medicine, Baskent University, Ankara, Turkey

**Keywords:** Key words Mitral stenosis, Echocardiography, Transcatheter valve interventions

## Abstract

**Background:** Spontaneous echo contrast(SEC) is an echocardiographic finding particularly found in left atrium of patients with mitral stenosis and known as a risk factor for stroke. However, its pathophysiology is not fully understood.

**Material and Method:** Forty-eight patients with mitral stenosis scheluded for percutaneous mitral valvuloplasty were included in the study. SEC was graded from 0 to 4 according to its density. Blood samples were taken from the aorta and left atrium during the procedure. Whole blood viscosity, plasma viscosity(PV) and peripheral blood smears were obtained and analysed separately from these sites. Prior to the procedure, all participants also underwent transthoracic and transesophageal echocardiography.

**Results:** Severe SEC(grade 3-4) was found in 23 patients(48%), remaining 25 patients(52%) had mild to moderate SEC(grade 0-1-2). The patients with atrial fibrillation(AF) and hypertension(HT) had more significantly severe SEC compared to other patients. Compared to patients with mild to moderate SEC, patients with severe SEC had increased age, body mass index, left atrial diameter, left atrial area and left atrial PV. However, ejection fraction, left atrial appendage(LAA) filling and emptying velocities, LAA lateral wall late systolic velocity, LAA fractional area change and pulmonary vein(PVe) systolic velocity were found to be significantly reduced in patients with severe SEC compared to mild to moderate SEC. On multiple linear regression analysis, AF, left atrium PV and left atrial diameter were the strongly correlated with SEC grade.

**Conclusion:** We have shown that AF, HT, systolic dysfunction of LAA, increased left atrial dimensions, reduced ejection fraction, decreased PVe flow velocity and increased left atrial PV were related with the development of SEC in patients with mitral stenosis.

## INTRODUCTION

Mitral stenosis (MS) represents a significant health problem in the developing world. While the reported incidence of MS in the North America and Europe is approximately 1/100.000, this figure is almost 35/100.000 in Africa (1). The most frequent cause of MS is the rheumatoid involvement of the mitral valve due to rheumatic fever. MS is associated with an increased likelihood of thromboembolic as well as cerebrovascular events (2). Although the risk is more marked in the presence of concomitant atrial fibrillation (AF), such events may occur even in MS patients with sinus rhythm.

Spontaneous echo contrast (SEC) is a smoke-like echocardiographic appearance of the blood due to spontaneous acquisition of echogenicity by the blood flow, which displays no echocardiographic contrast under normal conditions. SEC is a significant predictor of thromboembolic events and is considered an independent risk factor for CVE in MS patients (3). Despite studies investigating the etiology of SEC, exact mechanisms of pathogenesis have not been elucidated.

Several physiological parameters (e.g. pressure, lumen diameter, vessel compliance, and peripheral vascular resistance) affect the blood flow in the vessels, although blood viscosity represents the single most important determinant of blood flow (4). Blood viscosity is a measure of the resistance of blood against flow, and is approximately 1.8 fold higher as compared to water. Complete blood viscosity has four distinct components: plasma viscosity (PV), hematocrit, aggregation and deformability of erythrocytes, and temperature (5). Patients with cardiovascular events or stroke have been found to have elevated blood viscosity as compared to normal controls (6). Although studies examining the association between SEC and the rheological characteristics of blood have been conducted, the characteristics and importance of the relationship between increased blood viscosity and SEC occurring in the left atrium remain largely unknown.

This study was undertaken with the objectives of assessing and comparing the blood viscosity, plasma viscosity, and peripheral blood smear findings in left atrial and aortic blood samples from patients undergoing percutaneous mitral valvuloplasty due to MS. Also the association of these parameters with the degree of left atrial SEC was examined, and the potential effects of the morphological and functional characteristics of the left atrium (LA) and left atrial appendix (LAA) as well as of the pulmonary venous flow on the degree of SEC were investigated.

## MATERIALS AND METHODS

A total of 48 MS patients undergoing percutaneous mitral valvuloplasty between January 2016 and December 2018 at the Department of Cardiology, Adana Dr. Turgut Noyan Application and Research Center, Başkent University were included. The study protocol was approved by the Ethics Committee for Clinical Research, Başkent University (approval no: KA16/155). All patients provided written informed consent before participation in the study.

This research was done without patient involvement. Patients were not invited to comment on the study design and were not consulted to develop patient relevant outcomes or interpret the results. Patients were not invited to contribute to the writing or editing of this document for readability or accuracy.

Patients diagnosed with moderate to severe MS based on echocardiographic findings (mitral surface area < 1.5 cm2 and/or mean gradient > 10 mmHg) and scheduled for percutaneous mitral valvuloplasty were considered eligible. Those who had CVE in the past month were excluded as were those with thrombus in LAA or an ejection fraction below 45%.

Prior to the procedure, all participants underwent transthoracic and transesophageal echocardiography to assess the mechanical functions of the left ventricle, structure and the functions of the mitral valve, morphology and functions of the left atrium and left atrial appendix, and pulmonary venous flow. Echocardiography was performed using the imaging techniques, as described in the European Association of Echocardiography guidelines.

Transthoracic echocardiographic (TTE) examinations (2-dimensional, M-mode, color Doppler echocardiography) were carried out at parasternal and apical windows after at least 15 minutes of rest and in the left lateral position in each patient, using a Philips Epiq 7C device and X5-1 probe (1-5 MHz). MKA was calculated using the planimetric method in the short axis. The left atrial surface area and diameter, diameter of the right cardiac chambers, left atrial wall velocity, isovolumic relaxation time (IVRT), mitral E wave deceleration time (MSZ), TAİZ, left ventricular beat ratio (SVAO), and the septal and lateral wall DDE values of the mitral annulus were measured. Then, 10% lidocaine spray was used to provide local anesthesia of the pharynx, and patients were sedated with midazolam i.v. (2-6 mg). A TEE was performed using a Philips Epiq 7C device and X7-2t (2-7 Mhz) probe. After standard images of the cardiac chambers and valves were obtained, left atrium and left atrial appendix were examined carefully for the presence of SEC and thrombus. The severity, intensity, mobility, and localization of the SEC were graded in a scale from 0 to 4 by consensus readings of two senior echocardiographers blinded to study patients using the following criteria (Table 1) (7). LAA area, wall velocity, flow rates, and PVe flow were recorded. When calculating the LAA area, the area below the line drawn between limbus and aorta was taken into consideration. LAA flow rates were measured 2 mm below the LAA outlet, while wall velocities were measured from the mid-portion of the outer and inner walls. PVe flow was determined by placing the pulse Doppler cursor 5 to 10 mm into the left atrial orifice of the left superior pulmonary vein.

**Table 1.** SEC Definiton and Grading.

| SEC Grade | Definition |
| --- | --- |
| 0 | no SEC |
| 1 | Mild (minimal echogenicity in LAA, or rarely within the left atrium. Appears transiently during the cardiac cycle; using the “gain” settings, it may appear and disappear). |
| 2 | Mild to Moderate (Same distribution as in 1, but more intense. May be detected without changing “gain” settings). |
| 3 | Moderate (LAA displays intense sinuosity and waves, with lesser intensity in the left atrium. Despite fluctuations during the cardiac cycle, can be permanently detected |
| 4 | Severe (LAA has an echodense appearance with very slow sinuosity, and generally has the same intensity in the left atrium). |

SEC: Spontaneous echo contrast, LAA: Left atrial appendage

During PMBV, vascular sheaths have been placed onto the femoral vein and artery. Firstly, 15 ml of blood was obtained from the descending aorta using a pigtail catheter. After septostomy, 15 ml of blood was obtained from the left atrium, followed by the administration of unfractionated heparin. These two blood samples were placed in two 13 × 75 BD Vacutainer glass K3EDTA tubes of 2 ml with purple caps. Whole blood viscosity and plasma viscosity were assessed in these blood samples and were measured at 37 C using a Brookfield rotational mechanical viscositometer device (DV II + Pro, Brookfield Engineering Labs, MA, USA). Peripheral blood smears were prepared from bloods in EDTA tubes. After staining the peripheral smears with May-Grünwald Giemsa stain, the samples were examined by a hematology specialist blinded to patients. Differences in erythrocyte morphology, rouleau formation, and platelet aggregates were recorded.

For statistical analyses, SPSS 17 for Windows (Statistical Program for the Social Services Inc., Chicago, IL, USA) software pack was used. Continuous variables were expressed as mean ± standard deviation. Normal distribution of the variables was tested with Kolmogorov Smirnov test. Continuous variables were compared with independent sample T test. For data without normal distribution, non-parametric Wilcoxon-Signed Rank test was used for the comparison of two dependent groups, and Mann-Whitney U test for independent two groups. Chi-square test was used to assess discrete variables. A p value of less than 0.05 was considered statistically significant.

## RESULTS

The mean age of the 48 participants was 44.88 ± 14.09 years, while the mean body mass index was 28.53 ± 6.45 kg/m2. Most participants (85.4%) were female. Table 2 shows the general clinical and demographic characteristics of the patients.

**Table 2.**
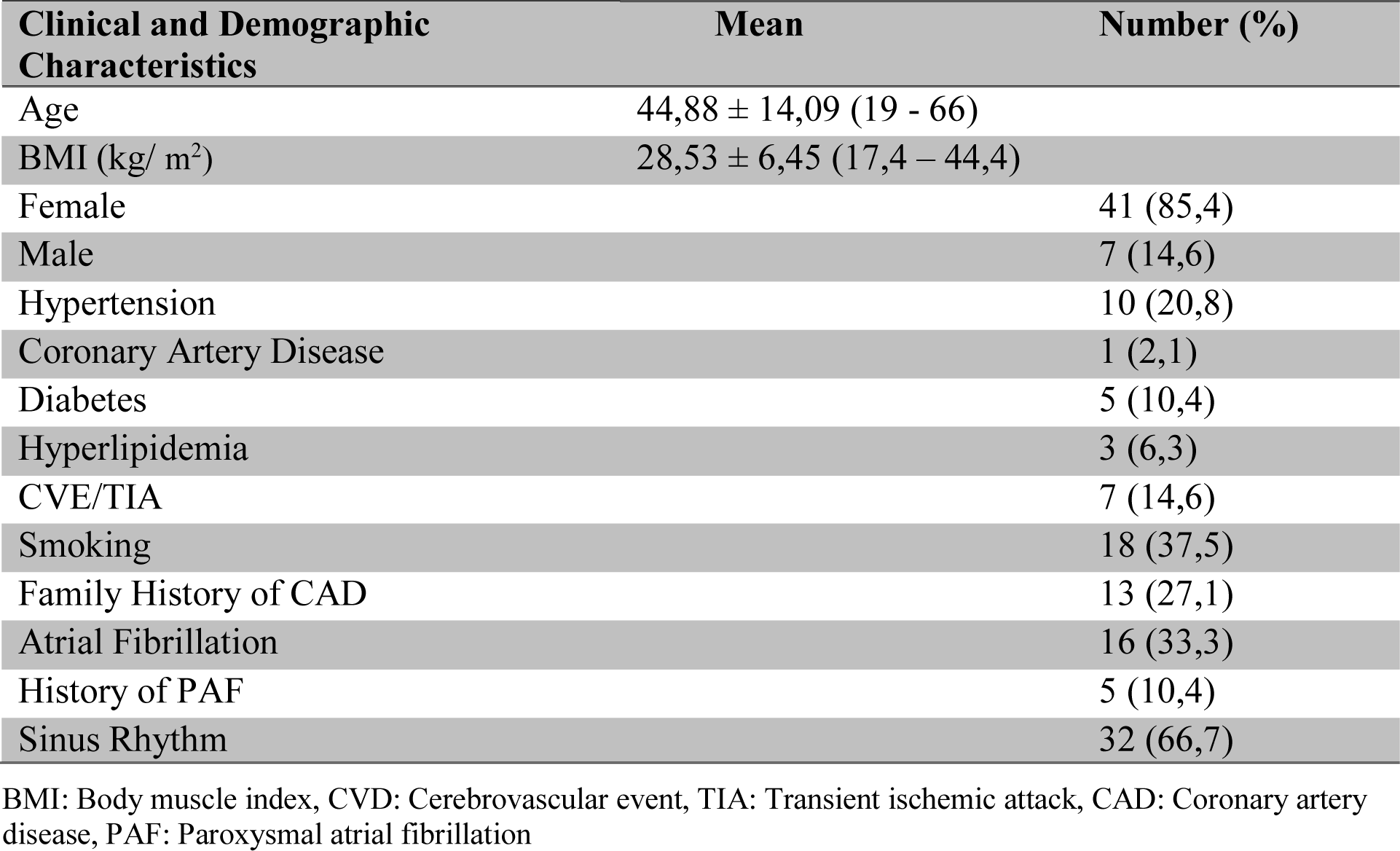
Clinical and Demographic Characteristics.

For certain statistical analyses, patients were divided into two groups as those with “mild to moderate SEC” (a SEC grade 0, 1, or 2) or those with “severe SEC” (a SEC grade of 3 or 4). Accordingly, 25 patients (52%) had mild to moderate, and 23 patients (48%) had severe SEC, respectively. The mean age and VKİin the latter group were significantly higher (p < 0.001 and p=0.002, respectively). There were no gender differences with respect to SEC severity (p=0.47).

Although more intense SEC images were observed in the left atrium an patients with a history of CVE or transient ischemic attack (TIA) than those without these conditions, the difference was not statistically significant (p=0.075). Cigarette smoking also did not affect the SEC severity significantly (p=0.258). On the other hand, patients with AF had significantly more severe SEC in their left atria (p=0.001). Five (16%) of the 32 patients in sinus rhythm had a known history of paroxysmal atrial fibrillation (PAF). Again, a history of PAF did not show any significant correlations with the severity of SEC (p=0.568).

Table 3 summarizes the two-dimensional TTE and Doppler results in patient groups. These data showed a significant association of SEC severity with left atrial, right atrial as well as right ventricular diameters, and with the length of the left atrium. Also, significant associations between SEC and the increase in left ventricular end systolic volume and the decrease in left ventricular ejection fraction have been found. Also, patients with severe SEC had significantly increased left atrial surface measurements obtained in two- and four-chamber windows at different phases of the cardiac cycle.

**Table 3.** Two-dimensional TTE and Doppler Results.

|  | Mild to Moderate SEC (0 - 1- 2) | Severe SEC (3-4) | p value |
| --- | --- | --- | --- |
| Left atrium (cm) | 4,74 ± 0,52 | 5,35 ± 0,71 | 0,001 |
| Right atrium (cm) | 3,18 ± 0,44 | 4,01 ± 0,63 | < 0,001 |
| Right ventricle (cm) | 2,60 ± 0,38 | 3,05 ± 0,56 | 0,002 |
| Left atrium length (cm) | 6,73 ± 0,76 | 7,52 ± 1,24 | 0,010 |
| End diastolic volume (ml) | 82 ± 24 | 87 ± 18 | 0,430 |
| End systolic volume (ml) | 32 ± 12 | 39 ± 12 | 0,037 |
| Left ventricular EF (%) | 61 ± 6 | 56 ± 8 | 0,024 |
| LA end systolic area (4 chamber)(cm<sup>2</sup>) | 28,9 ± 5,9 | 37,3 ± 10 | 0,001 |
| LA end diastolic area (2chamber)(cm<sup>2</sup>) | 25,7 ± 6,6 | 33,8 ± 10 | 0,002 |
| LA end systolic area (4 chamber)(cm<sup>2</sup>) | 25,9 ± 5,5 | 34,9 ± 9,5 | < 0,001 |
| LA end diastolic area (2 chamber)(cm<sup>2</sup>) | 22,7 ± 6,1 | 31,7 ± 9,1 | < 0,001 |
| Mitral deceleration time | 561 ± 195 | 560 ± 171 | 0,980 |
| Isovolemic relaxation time (msn) | 71 ± 20 | 80 ± 17 | 0,094 |
| Mitral E wave (cm/sec) | 225 ± 43 | 226 ± 39 | 0,940 |
| Mitral valve gradient (mmHg) | 13,6 ± 4,1 | 11,9 ± 3,9 | 0,059 |
| Mitral valve area (cm<sup>2</sup>) | 1,10 ± 0,20 | 1,13 ± 0,21 | 0,580 |
| Wilkins Score | 7,4 ± 0,7 | 7,6 ± 1,1 | 0,443 |
| Mitral annular mean E' | 6,7 ± 2,5 | 6,9 ± 1,9 | 0,686 |
| Mitral annular septal A' | 6,7 ± 2 | 6,1 ± 1,8 | 0,518 |
| Mitral annular septal S' | 8,2 ± 1,5 | 7,6 ± 2,1 | 0,221 |
| Mitral annular lateral A' | 8,6 ± 2,8 | 6,6 ± 2 | 0,076 |
| Mitral annular lateral S' | 8,7 ± 1,2 | 7,9 ± 2,3 | 0,153 |

EF: Ejection fraction, LA: Left atrium, MDT: Mitral E wave deceleration time, E: Early diastolic mitral inflow velocity, E': Early diastolic mitral annular velocity, A': Late diastolic mitral annular velocity, S': Systolic mitral annular velocity

After 32 patients with sinus rhythm were divided into two groups based on the severity of SEC, left atrial mean wall velocity and total atrial conduction time (TACT) were analyzed to compare these two groups with respect to left atrial functions (Table 4). Although increasing severity of SEC was positively correlated with TAİZ (r=363, p=0,044) and negatively correlated with LA anterior wall mean velocity (r=-357, p=0.045), the difference in these two parameters between the two groups defined on the basis of SEC severity was not significant. Again, other echocardiographic and Doppler parameters of TTE did not show significant associations with SEC severity.

**Table 4.** Distributions of LA Mean Wall Velocity and TACT According to Severity of SEC in The Patients With Sinus Rhythm.

|  | **Mild to Moderate SEC (0 - 1- 2)** | **Severe SEC (3-4)** | **p value** |
| --- | --- | --- | --- |
| IAS mean velocity (cm/sec) | 9,8 ± 4,8 | 8,0 ± 3,1 | 0,367 |
| LA lateral wall mean velocity (cm/sec) | 10,3 ± 3,5 | 8 ,4± 3,9 | 0,322 |
| LA anterior wall mean velocity (cm/sec) | 10,3 ± 4 | 7,4 ± 2,8 | 0,085 |
| LA inferior wall mean velocity (cm/sec) | 9,7 ± 2,7 | 8,8 ± 3,2 | 0,461 |
| TACT (msn) | 132 ± 30 | 149 ± 12 | 0,200 |

IAS: Inter atrial septum, LA: Left atrium, TACT: Total atrial conduction time

TEE findings regarding LAA morphology and function as well as the left upper PVe flow were examined in detail (Table 5). In patients with severe SEC, LAA filling and emptying were significantly lower as compared to the other group. Individual comparisons of inner and outer LAA wall tissue Doppler velocities between the two groups showed a decrease in all four parameters in the severe SEC group, although the only significant difference was noted for LAA outer wall systolic velocity (p=0.002). Comparison of the maximum and minimum LAA areas between the two groups did not reveal any differences. However, the fractional surface area alteration was significantly lower in the severe SEC group vs. other patients (21.3 ± 8.8% vs. 30.6 ± 10.2%, respectively; p=0.001).An examination of the difference in pulmonary venous blood flow rate showed lower systolic, diastolic, and atrial systolic reversal wave in the pulmonary vein among patients with severe SEC, although the only significant difference was identified in PVe systolic velocity (p=0.022).

**Table 5.** TEE Findings of Left Atrial Appendage and Pulmonary Vein.

|  | **Mild to Moderate SEC (0 - 1- 2)** | **Severe SEC (3-4)** | **p value** |
| --- | --- | --- | --- |
| LAA filling velocity(cm/sec) | 38,7 ± 13,4 | 26,9 ± 8,3 | 0,001 |
| LAA emptying velocity (cm/sec) | 36,6 ± 13,6 | 25,7 ± 9,1 | 0,002 |
| LAALW late systolic velocity (cm/sec) | 7,2 ± 2,4 | 5,3 ± 1,5 | 0,002 |
| LAALW late diastolic velocity (cm/sec) | 7,0 ± 3,7 | 6,0 ± 1,7 | 0,243 |
| LAAMW late systolic velocity (cm/sec) | 7,7 ± 2,9 | 6,4 ± 1,7 | 0,055 |
| LAAMW late diastolic velocity (cm/sec) | 7,2 ± 3,0 | 6,9 ± 2,3 | 0,288 |
| LAA largest area (cm<sup>2</sup>) | 5,27 ± 1,30 | 5,16 ± 1,41 | 0,770 |
| LAA smallest area (cm<sup>2</sup>) | 3,70 ± 1,21 | 4,05 ± 1,18 | 0,316 |
| LAA fractional area change (%) | 30,6 ± 10,2 | 21,3 ± 8,8 | 0,001 |
| PVe systolic wave (cm/sec) | 51,1 ± 16,0 | 40,6 ± 14,7 | 0,022 |
| PVe diastolic wave (cm/sec) | 38,1 ± 12,3 | 33,4 ± 13,2 | 0,209 |
| PVe atrial systolic reversal wave (cm/sec) | 32,2 ± 12,4 | 28,2 ± 8,6 | 0,202 |

LAA: Left atrial appendage, LAALW: Left atrial appendage lateral wall, LAAMW: Left atrial appendage medial wall, PVe: Pulmonary vein,

Table 6 summarizes the viscosity in blood samples obtained from the left atria and aorta of the study participants. A positive correlation was found between increasing SEC severity and left atrial PV and aortic PV (r=297, p=0.040; r=291, p=0.045, respectively). Patients with severe SEC also had higher whole blood and plasma viscosity in left atrial and aortic samples, although the only significant difference was found in left atrial PV (p=0.037).

**Table 6.** The Viscosity in Blood Samples Obtained From the Left Atria and Aorta.

|  | Mild to Moderate SEC<br>(0 - 1- 2) | Severe SEC<br>(3-4) | p value |
| --- | --- | --- | --- |
| Left atrium WBV (mPa-s) | 4,10 ± 0,69 | 4,20 ± 0,51 | 0,552 |
| Left atrium PV (mPa-s) | 1,69 ± 0,22 | 1,86 ± 0,33 | 0,037 |
| AorticWBV (mPa-s) | 3,94 ± 0,62 | 4,07 ± 0,48 | 0,470 |
| Aortic PV (mPa-s) | 1,55 ± 0,26 | 1,69 ± 0,24 | 0,059 |

WBV: whole blood viscosity, PV: Plasma viscosity,

The comparison of aortic and left atrial PV according to left atrial SEC severity and rhythm is shown in Figure 1. As long as the overall group of participants were considered regardless of SEC severity and rhythm status, left atrial PV values were significantly higher as compared to aortic PV (p=0.001). A comparison of left atrial and aortic blood values between the groups defined on the basis of SEC severity showed significant elevation in favor of left atrial PV in both groups (p=0.025 and p=0.022, for severe and mild to moderate SEC, respectively).

**Figure 1.**
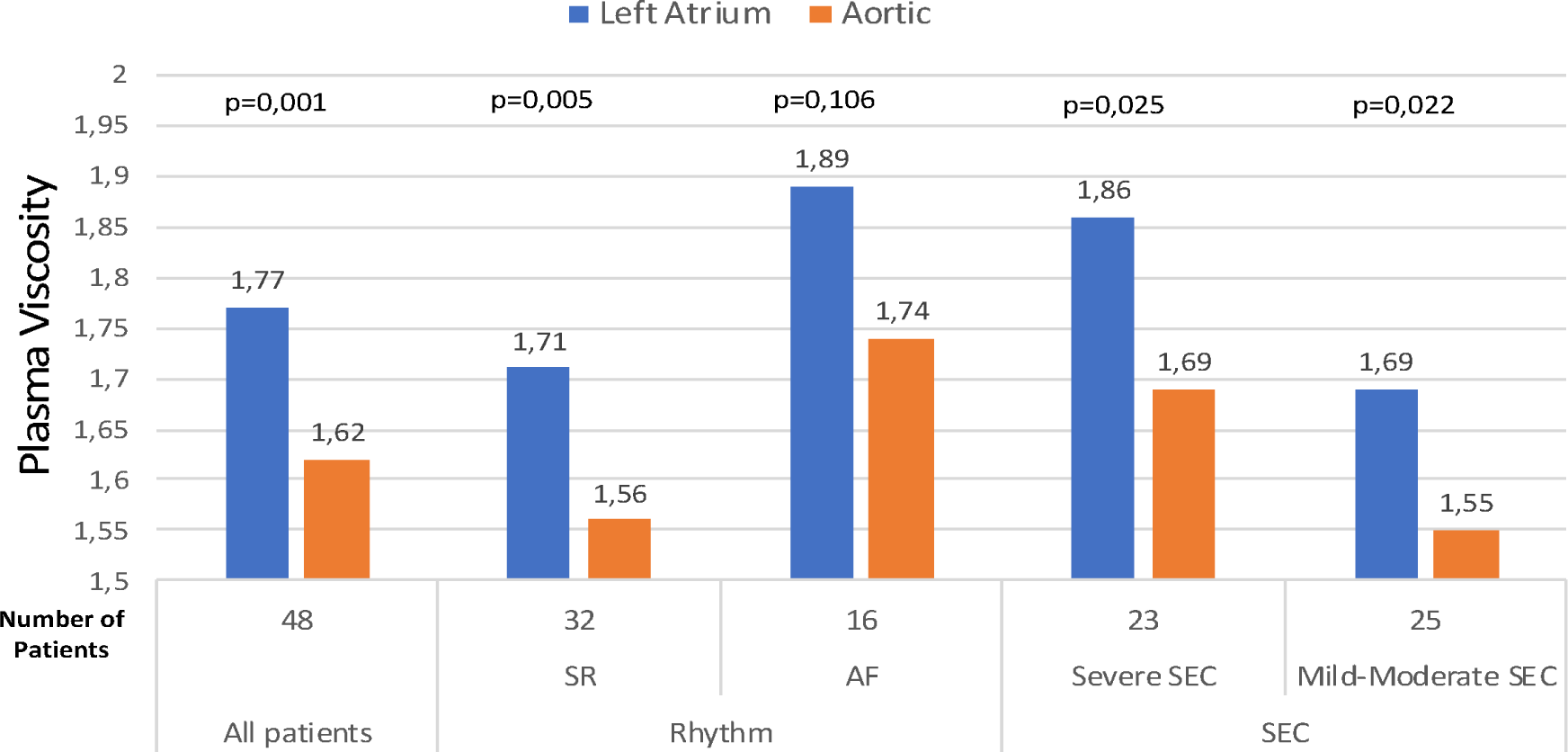
The comparison of aortic and left atrial plasma viscosity according to left atrial spontaneous echo contrast (SEC) severity and rhythm.

Comparison of viscosity results in terms of sinus rhythm status showed no differences between left atrial and aortic PV in patients with AF, while in those with sinus rhythm, significantly higher left atrial PV was found irrespective of SEC severity (p=0.005). Left atrial WBV and aortic WBV did not exhibit any significant differences when patient sub-groups were compared.

## DISCUSSION

Previous studies showed an increased risk of thrombosis in MS patients that is independent of the presence of AF, with higher incidence of CVE than those without MS (2). Furthermore, studies showing an association of SEC severity with CVE and thrombus formation in the LAA suggest that SEC may represent a potential causative factor in these conditions (3). The relationship between SEC and left atrial as well as LAA diameter and functions has been firmly established (8). Also, the link between several biomarkers and SEC has also been examined (9). Despite these various studies, the exact mechanisms of SEC have not been fully elucidated. In the current study, the left atrial and aortic blood viscosity and peripheral smears, morphology and function of the LAA and left atrium, and pulmonary vein flow patterns were examined in detail, in an effort to shed a light on the etiology of SEC.

Approximately one third of our participants had AF, all of whom had severe (grade 3 or 4) SEC. A multi-variant linear regression analysis revealed that the left atrial PV and left atrial diameter as well as the presence of AF emerged as the most important independent determinants of the severity of SEC. In a TEE study by Kewal C. et al. (10) involving 200 MS patients, SEC was found to be closely linked with AF, consistent with our observations.

Sadanandan et al. (11) reported a significant association between left atrial SEC and a history of CVE in a cohort of patients with sinus rhythm. In our study, although patients with a history of CVE/TIA had more severe SEC, the difference was not significant. In the abovementioned study, patients with SEC were compared with controls without SEC. On the other hand, absence of a control group and assignment of our patients based on the severity of SEC may probably explain the absence of a similar significant difference.

A higher BMI and obesity are associated with an elevated risk of AF. Also, obesity represents a risk factor for ischemic CVE, thromboembolic events, and death in patients with AF (12). A linear multivariate regression analysis of our patients showed that SEC severity was not only associated with the left atrial diameter and PV, but also with BMI, with increasing BMI values being more closely related to SEC severity. This observation is of particular note, since it shows the association between obesity and increasing severity of SEC, and hence of increased risk of thromboembolism, in a manner that is independent of the presence of AF. Also patients with severe SEC were older than those with mild to moderate SEC. This last observation is consistent with the findings of Kewal C. et al. (10).

Multivariate linear regression analysis also showed that the left atrial diameter was one of the significant independent predictors of SEC severity. Similarly, Soydaş C. et al. (13) also observed an association between SEC and increasing dimensions of LA. Reduced left ventricular ejection fraction also has been reported to be correlated with left atrial SEC severity (14, 15, 16). In line with such reports, a reduced left ventricular beat rate was associated with increasing SEC severity in this cohort. This observation, also corroborated with our findings, probably results from the increased stasis of the blood in the left atrium due to left ventricular dysfunction.

Previous studies showed that DDA data obtained from LA walls, is an important parameter reflecting the regional atrial function (17, 18, 19). To the best of our knowledge, our study represents the first of its kind, as it compares the wall velocities that are obtained with DDE and that directly mirror SEC and LA functions. Although patients with severe SEC had lower overall LA wall velocity, the difference did not reach statistical significantly, as also was the case for TACT. These parameters were only measured in patients with sinus rhythm, since they are directly related to the P wave and atrial contraction. Presence of AF in the majority of our sample may account for our failure to detect significance.

The association between LAA dysfunction and SEC severity and thromboembolic events has been well established (20). Reddy VG et al. (21) showed that MS patients with sinus rhythm had significant improvement of LAA flow velocities early after PMBV and that the degree of improvement was related with the reduction in the severity of SEC. Another echo parameter providing information on LAA functions is the LAA wall velocities measured using the tissue Doppler technique. In AF and MS patients, LAA wall velocities measured by DDE are lower as compared to healthy controls (22, 23, 24). Again, in another previous study, severity of SEC was found to be negatively correlated with the late systolic wall velocity in the LAA in patients with MS (25, 26). In line with these reports, we also confirmed the association of LAA flow velocity as well as the reduced late systolic velocity of the outer LAA wall and increasing SEC severity in MS patients. LAA fractional surface area change, another indicator of LAA functions, was lower in patients with severe SEC, in parallel with previous observations (27). Our results seem to support the close association of severe SEC with LAA functions and morphology.

In 2002, Bollmann A. et al. (28) described the reduced PVe flow and the presence of SEC for the first time. In 2005, Donal E. et al. (29) showed that systolic PVe flow was a marker for the improvement of left atrial functions, in patients undergoing pulmonary vein isolation with ablation. Similar to that study, we also observed significantly reduced PVe flow in patients with severe SEC. While this association was described in patients with AF in the study by Bollman A. et al., in our study such a relationship was for the first time shown in MS patients irrespective of the presence of AF. However, this finding should be cautiously interpreted, as one third of our patients had AF rhythm.

Theoretical explanations put forward for the mechanisms of SEC include erythrocyte aggregation, increased blood viscosity, or decreased shear stress. In a study by Roger E. Peverill at al. (30) involving a group of MS patients with sinus rhythm, the severity of SEC correlated with hematocrit value and erythrocyte concentration. Again, Zotz RJ. Et al. (31) found higher platelet activation in left atrial blood samples from SEC patients as compared to that in the arterial blood and right atrial blood, suggesting that platelet aggregation may play a role in the pathogenesis of SEC, similar to the assertions of Wang J. et al. (32). In one study by Atak R. et al. (33) MS patients with sinus rhythm were found to have elevated left atrial coagulation activity, independent of the presence of SEC. In one study involving 50 patients with a history of CVE, Briley DP et al. (34) observed an association between plasma viscosity and SEC. In the light of such previous publications, we also aimed at determining the role of blood viscosity in the occurrence of SEC by also examining the left atrial blood samples.

Although a significant positive correlation between SEC severity and both aortic PV and left atrial PV was found, a comparison of the study groups showed a significant increase in left atrial PV only among those subjects with severe SEC. Again, independent of the severity of SEC, MS patients had significantly higher left atrial PV values than aortic PV values. These results seem to support our hypothesis that increased local plasma viscosity in the left atrium may play a significant causative role in the occurrence of SEC. Normal plasma viscosity at 37 □ is 1.10 to 1.35 mPa/sec (milli-Pascal/second) independent of the age and gender (35). In our study, PV in both aortic and left atrial blood samples was significantly higher as compared to these normal values. However, one should not jump into the conclusion that MS patients had elevated PV values based on these findings, as the study included no control group by design. Although our patients with more severe SEC had increased rouleaux formation in their aortic and left atrial peripheral smears, this increase was insignificant. Again, peripheral smears from two patient groups did not exhibit significant differences with regard to platelet aggregation or distribution.

## STUDY LIMITATIONS

One of the limitations of our study is the small number of study subjects, and the results may not be generalizable to larger populations. It is currently unclear whether there is a strong relationship between plasma viscosity and SEC. Thus, further prospective studies with a larger population are needed to address this question.

## CONCLUSION

Our results showed an association between development of SEC and AF, LAA systolic dysfunction, increased left atrial dimensions, reduced left ventricular ejection fraction, reduced PVe flow rate, and increased left atrial plasma viscosity in patients with mitral stenosis. The observed association between plasma viscosity and SEC in this study may represent a novel mechanism for the possible etiology of SEC that warrants further studies.

## Data Availability

Data are available upon reasonable request

